# Integrity of dopaminergic terminals in the caudate nucleus is relevant for rest tremor in Parkinson’s disease

**DOI:** 10.1101/2024.04.04.24305353

**Authors:** Marcelo D. Mendonça, Pedro Ferreira, Francisco Oliveira, Raquel Barbosa, Bruna Meira, Durval Costa, Albino Oliveira-Maia, Joaquim Alves da Silva

## Abstract

Resting tremor (RT) is a Parkinson’s disease (PD) symptom whose relationship with the dopaminergic system remains puzzling. To clarify this relation, we analysed data from 432 subjects from the PPMI (Parkinson’s Progression Markers Initiative) database, a second cohort of 57 PD patients and controls and a third cohort of 86 subjects referred for dopamine transporter single-photon emission computed tomography (DaT-SPECT). Caudate, but not putamen binding potential (CBP, PBP) was higher in patients with RT. The presence of a higher CBP at the baseline and a relative sparing of CBP were related to the development of RT in the 2^nd^ year. In the smaller cohorts, a 4-6 Hz oscillation-based metric extracted from inertial sensors correlated with RT amplitude and distinguished groups with DA denervation from controls. The same metric was correlated with CBP, but not PBP in subjects with a denervated DaT-SPECT. *In silico* modelling uncovered that higher CBP in patients with RT was fundamental to explain the associations and correlations between RT and DaT-SPECT described in our and other datasets. These results support that the integrity of caudate dopaminergic terminals could be relevant for RT pathophysiology. Unbiased assessment methods, such as inertial sensors, can be important to understand PD pathophysiology.

## Introduction

Tremor, one of the main motor symptoms of Parkinson’s disease (PD) is classically seen at rest and with a frequency of 4-6 Hz(1). Tremor is a highly heterogeneous symptom: It is not present in all patients, it may occur in different body areas, it may range in amplitude, and it is vulnerable to environmental stressors(1,2). Paired with this rich phenomenology, the pathophysiology of tremor is distinct from other PD motor symptoms and remains challenging to understand. Dopamine (DA) loss is necessary for rest tremor (RT) to be present(3,4), however, it is well known that an important number of PD patients present RT with no response to dopamine replacement therapy(5) or even worsening of tremor(6,7). This suggests that the link between dopamine depletion and RT is more complex than a simple DA dose dependency model.

Neuropathological studies pointed out that in PD patients with RT, there is a higher loss of A8 dopaminergic neurons (retrorubral area, RRA)(8). A similar observation was found when comparing the phenotypic heterogeneity of RT in non-human primate models of PD(9–11). RRA neurons were documented to project to the globus pallidus (GP)(12,13), which emerged as an anatomical(14) and functionally relevant region in RT(15). This node has been proposed as a switch in the well-established “dimmer-switch hypothesis” for RT(16). According to this hypothesis, DA loss in the RRA projections to the GP initiates tremor-related oscillations in the basal ganglia circuits that are propagated via motor cortex to the cerebello-thalamo-cortical circuits(17,18). The cerebello-thalamic circuit, involving the thalamic ventral intermediate nucleus (VIM) acts as a dimmer that maintains and amplifies the tremor. This is in line with the well-known ability of VIM deep brain stimulation (DBS) to decrease the activity in the thalamo-cortical circuit and reduce tremor(19,20) with minimal effect on bradykinesia or rigidity(21). However, evidence on the relationship between pallidal DA and tremor remains confusing as this model is not aligned with observations that tremor dominant forms present higher pallidal dopamine transporters (DaT) binding(18) and PD patients with tremor present relative pallidal DaT sparing(17).

Uncertainty regarding a potential differential loss of dopamine neural terminals between patients with tremor-dominant and non-tremor-dominant forms of PD is not only circumscribed to pallidal dopamine. Molecular imaging studies using ^123^I-FP-CIT (a radiopharmaceutical that binds to DaT) have shown heterogeneous results with some authors reporting higher(22–24) ^123^I-FP-CIT binding in the caudate and putamen(22,23,25) of tremor patients, other studies reporting no difference in caudate(25,26) or putamen(26,27) or even a reduced caudate binding potential in tremor-dominant PD patients(27). Heterogeneity could emerge from small datasets with retrospective clinical assessments, and heterogeneous groups of tremor-dominant patients, making relevant an assessment that is symptom-driven(28–30). Classical phenotypical classification of PD patients in tremor-dominant or postural instability and gait difficulty phenotypes consider, not the presence of the symptom *per se*, but its relative severity in comparison to other motor symptoms. If we consider that specific symptoms emerge from specific brain networks dysfunction, their presence (more than relative severity) should be the focus to reduce heterogeneity in findings.

Here, we take advantage of clinical and DaT imaging datasets, enriched with inertial sensors assessment, to clarify the role of caudate and putamen dopaminergic innervation in RT pathophysiology. We expand on current RT models and demonstrate that the integrity of caudate dopaminergic terminals is related to the presence and emergence of RT, in line with pre-clinical evidence that manipulations of caudate cholinergic or dopaminergic systems lead to the development of tremor(31–33).

## Results

### Presence of RT is associated with higher integrity of caudate dopaminergic terminals

We identified 432 patients with PD with a follow-up of at least 24 months since study inclusion and available information on RT at this timepoint (Fig. 1A). From these subjects, 66.4% presented RT at this evaluation. Patients with tremor were slightly older than patients without any tremor (62.8±0.6 vs. 60.3±0.8, *t*_289.1_=2.428, *P*=0.0158, Fig. 1B), and the groups had a comparable distribution regarding sex (% of females in tremor group: 40.42%, % of females in no-tremor Group: 34.48%, χ^2^=1.118, *df*=1, *P*=0.2904, Fig. 1C). Patients with RT, when compared with patients with no RT have a significantly higher ^123^I-FP-CIT CBP that does not differentially change between groups during a 2-year period (Repeated Measure Mixed-Effect Model: Clinical Group: *F*_1,420_=10.21, *P*=0.0015; Time: *F*_2,601_=166.3, *P*<0.0001; interaction: *F*_2,601_=0.7794, *P*=0.4591, year 0 least squares mean difference: 0.1431±0.0549, p=0.0093; year 1 least squares mean difference: 0.1788±0.0616 *P*=0.0075; year 2 least squares mean difference: 0.183±0.0559; *P*=0.0032 Fig. 1D). No significant group difference was seen in PBP (Repeated Measure Mixed-Effect Model: Clinical Group: *F*_1,420_=3.721, *P*=0.0544; Time: *F*_2,601_=122.1, *P*<0.0001; interaction: *F*_2,601_=0.01066, *P*=0.9894). Besides per definition clinical differences in RT (Fig 1E), there was a difference in action tremor scores (Fig 1F), but no relevant differences were found in bradykinesia (Fig 1G) or rigidity scores (Fig 1H). Yearly analysis that included brain region (caudate/putamen) as a variable disclosed an interaction between the region and RT, supporting a relation that is region-specific between CBP and RT (detailed analysis in Supplementary Figure 1).

**Figure 1.**
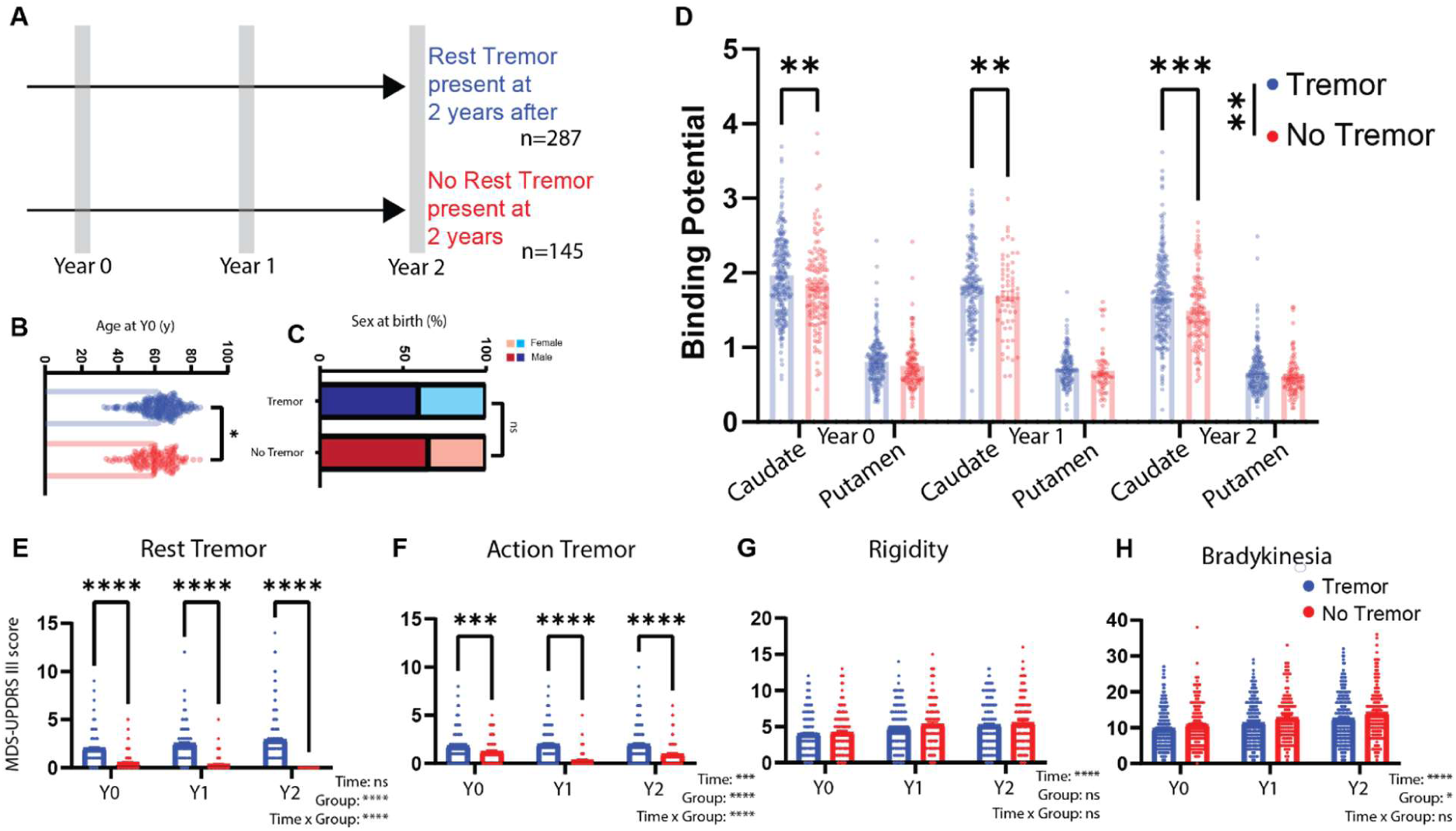
The presence of RT in PD is associated with higher caudate binding potential. **(A)** Patients included in the PPMI study with MDS-UPDRS III tremor data in off state, available at Year 2 after inclusion (n=432) were selected. Groups were defined by the presence of RT (Score on the MDS-UPDRS III 3.17 > 0) or its absence. Data on baseline (Year 0) and 12 months after (Year 1) was also collected. (**B)** Age at baseline in the Tremor and No-Tremor groups. (**C)** Sex at birth proportion in the Tremor and No-Tremor groups. (**D)** Caudate and putamen biding potential in the Tremor and No-Tremor groups at baseline and follow-up. (**E)** RT scores (MDS-UPDRS III 3.17 score). Time: *P*=0.0553, Group: *P*<0.0001, Time × Group: *P*<0.0001. Post-hoc (Šídák’s multiple comparisons test), *P*<0.0001 for all comparisons (**F)** Action tremor scores (MDS-UPDRS III 3.15 and 3.16 score). Time: *P*=0.0001, Group: *P*<0.0001, Time × Group: *P*<0.0001. Post-hoc (Šídák’s multiple comparisons test), *P*<0.001 for all comparisons (**G)** Rigidity scores (MDS-UPDRS III 3.3 score). Time: *P*<0.0001, Group: *P*=0.3361, Time × Group: *P*=0.7546. **H)** Bradykinesia scores (MDS-UPDRS III 3.4, 3.5, 3.6, 3.7, 3.8, 3.9 and 3.14 score). Time: *P*<0.0001, Group: *P*=0.0148, Time × Group: *P*=0.6441.

### The Integrity of caudate dopaminergic terminals is related to the development of RT

Patients with and without RT after the 2-year follow-up period could have had different symptomatic trajectories regarding tremor. To study the development of tremor as a symptom, we categorized each group based on the presence or absence of RT in the first clinical assessment (Fig. 2A, Supplementary Fig 2A).

**Figure 2.**
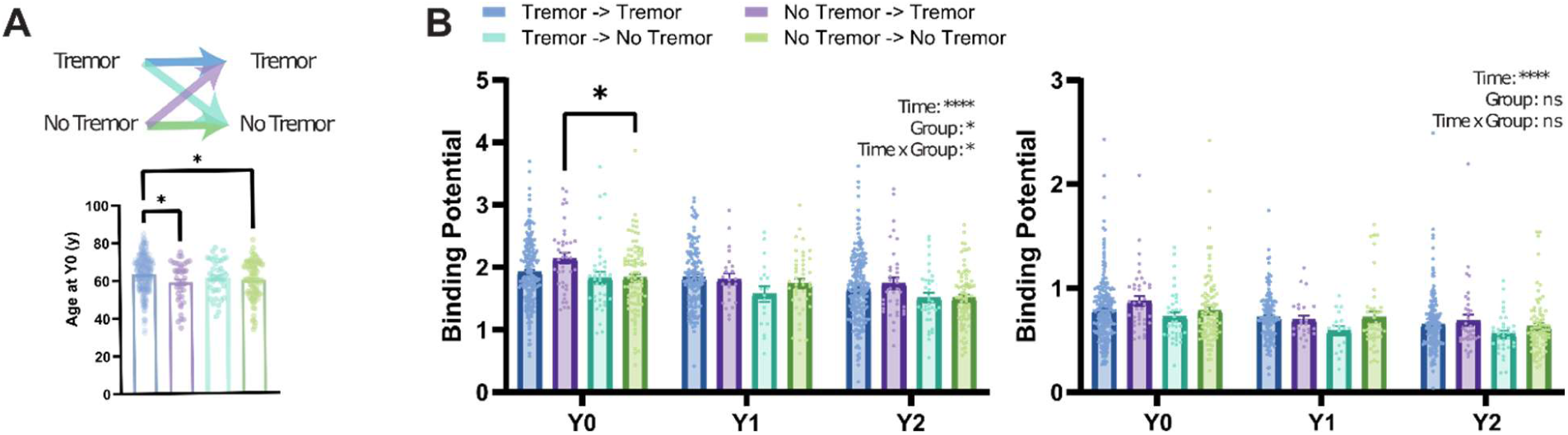
Integrity of caudate dopamine terminals is related to the development of RT. (A) Left: Patient groups were sorted by the baseline (Year 0) presence of tremor and included in four different groups depending on maintaining or switching their phenotype. Right: Age at inclusion for the different groups (*F*_3,405_=4.802 *P*=0.0027; Post-hoc comparisons (Holm-Šídák’s multiple comparisons test: Tremor->Tremor vs. No-tremor->Tremor (63.50±0.63 vs. 58.97±1.64, *P*=0.030), Tremor->Tremor vs. No-Tremor-> No-tremor (63.50±0.63 vs. 59.79±1.04, *P*=0.0133)). **(B)** Top: CBP at baseline and during the 2-year follow-up for the different groups. Bottom, PBP at baseline and during the 2-year follow-up for the different groups.

These four groups present significant differences in CBP (Repeated Measure Mixed-Effect Model: Clinical Group: *F*_3,396_=2.636, *P*=0.0494; Time: *F*_2,583_=122.3, *P*<0.0001; interaction: *F*_6,583_=2.2140, *P*=0.0473) with higher CBP in patients presenting without tremor that ended up developing RT when compared to those that did not develop it (year 0 least squares mean difference: 0.2878±0.099, *P*=0.0199, Fig 2B, top). CBP differences found in different tremor groups (Fig 2B) were not due to age (Fig 2B, bottom), rigidity (Supplementary Fig 2B) or bradykinesia (Supplementary Fig 2C).

Besides an expected reduction in binding with time, no significant differences were identified in the PBP (Repeated Measure Mixed-Effect Model: Clinical Group: *F*_3,396_=1.294, *P*=0.2761; Time: *F*_2,583_=93.34, *P*<0.0001; interaction: *F*_6,583_=0.6628, *P*=0.6628).

CBP at 2 years was associated with RT presentation at this timepoint (*OR* 7.235, 95% *CI*: 1.59– 39.06, *P*=0.0143, supplementary table 1). Nevertheless, if the integrity of caudate dopaminergic terminals is relevant for the development of RT, it would be expected that CBP already at baseline would inform the risk of RT at follow-up. In fact, a logistic regression showed that increased CBP at the baseline is associated with a significantly higher risk of developing RT even after adjusting for PBP, age, bradykinesia, and rigidity (*OR* 4.84, 95% *CI*: 1.558–16.93, *P*=0.0089, Table 1).

**Table 1.** Factors associated with development of RT. Logistic Regression with new onset of RT at year two as a dependent variable. Hosmer-Lemeshow test: *P*=0.230. AUC of the model: 0.647 (0.545–0.748; *P*=0.008).

|  | **OR** | **95% CI** | ***P*-value** |
| --- | --- | --- | --- |
| Intercept | 0.105 | 0.006 – 1.703 | 0.120 |
| **Caudate Y0** | **4.840** | **1.558 – 16.930** | **0.009** |
| Putamen Y0 | 0.305 | 0.040 – 1.935 | 0.222 |
| Age | 0.992 | 0.955 – 1.030 | 0.668 |
| Bradykinesia Y0 | 0.969 | 0.892 – 1.042 | 0.416 |
| Rigidity Y0 | 1.031 | 0.878 – 1.212 | 0.706 |

These results suggest that the relative sparing of caudate denervation in the context of progressive dopaminergic denervation is related to the development of RT in patients who did not present RT at the baseline, further supporting that the presence of RT is related with a more parsimonious loss of caudate dopaminergic terminals.

### Oscillations assessed with inertial sensors identify patients with DA depletion and correlate with tremor severity

Although the MDS-UPDRS is a valid and reliable instrument(34,35), the scoring has relevant within and between subject variability and some subscores of the resting tremor item are the less reliable of the whole scale(34). Inertial sensors have been successfully employed to quantify tremor in PD(36,37). Given the results showing an association between RT and CBP, we used inertial measurement units (IMUs) to further explore this relation using an objective and unbiased quantification of tremor.

We recruited 2 cohorts for this. One cohort was composed of patients with a clinical diagnosis of PD (n=28, Table 2) and healthy age-matched controls (n=29, Table 2). From now on this cohort will be called the PD/Control cohort. A second cohort was composed of patients that were referred for a DaT-SPECT at our clinical centre, as part of the diagnostic work-up of a movement disorder (n=86, Table 3). From now on this cohort will be called the DaT-SPECT cohort. In both cohorts, patients were assessed using the MDS-UPDRS and motion sensor data was collected while participants were standing still with their feet next to each other. For the second cohort, the evaluation was performed on the same day and before the SPECT image was collected, thus raters did not know, at the time of clinical assessment, if the participants had a positive or negative DaT-SPECT. Full cohort details can be found in the methods section.

**Table 2.** Hospital Egas Moniz (PD/control) cohort demographics.

|  | **PD** | **Control** | **P-value** |
| --- | --- | --- | --- |
| **n** | 28 (49.1%) | 29 (50.9%) |  |
| **Age (yr)** | 71.7 (10.7) | 69.4 (13.0) | 0.468 |
| **Female, n (%)** | 14 (50.0) | 16 (55.2) | 0.9 |
| **DA treatment, n (%)** | 22 (78.6) | - | - |
| **LEDD (mg/day)** | 389.0 (301.3) | - | - |
| **Symptom onset (months)** | 55.3 (53.6) | - | - |
| **MDS-UPDRS III Total** | 27.3 (13.1) | 2.5 (3.2) | <0.001* |
| **Rigidity** | 4.5 (3.3) | 0.2 (0.5) | <0.001* |
| **Bradykinesia** | 13.7 (8.4) | 1.1 (2.3) | <0.001* |
| **Rest tremor** | 1.8 (1.8) | 0.0 (0.2) | <0.001* |
| **Tremor** | 5.2 (4.6) | 0.6 (1.9) | <0.001* |

dopamine
LEDD – Levodopa Equivalent Daily Dose
MDS-UPDRS – Movement Disorder Society-sponsored Unified Parkinson's Disease Rating Scale

**Table 3.** Champalimaud Clinical Centre (DaT-SPECT) cohort demographics.

|  | **Positive** | **Negative** | **P-value** |
| --- | --- | --- | --- |
| **n** | 34 (39.5%) | 52 (60.5%) |  |
| **Age (yr)** | 65.1 (10.2) | 65.1 (14.9) | 0.988 |
| **Female, n (%)** | 21 (61.8) | 31 (60.8) | 1.000 |
| **DA treatment, n (%)** | 17 (50.0) | 8 (16.0) | 0.002* |
| **Tremor treatment, n (%)** | 1 (2.9) | 13 (26.0) | 0.013* |
| **LEDD (mg/day)** | 183.8 (295.2) | 51.6 (138.7) | 0.021* |
| **Symptom onset (months)** | 43.3 (70.1) | 41.2 (51.1) | 0.884 |
| **MDS-UPDRS III Total** | 21.4 (14.7) | 14.2 (12.2) | 0.021* |
| **Rigidity** | 2.9 (2.7) | 1.1 (1.6) | 0.001* |
| **Bradykinesia** | 11.3 (8.5) | 7.4 (7.2) | 0.031* |
| **Rest tremor** | 1.1 (1.7) | 0.7 (1.2) | 0.270 |
| **Tremor** | 3.2 (3.6) | 3.2 (4.0) | 0.976 |

DA – dopamine
LEDD – Levodopa Equivalent Daily Dose
MDS-UPDRS – Movement Disorder Society-sponsored Unified Parkinson's Disease Rating Scale

On the PD/Control cohort, per design, no differences were found in age or sex. As expected, PD patients had significantly higher bradykinesia, rigidity and RT (Table 2). Of the 86 participants in the DaT-SPECT cohort, 34 had evidence of dopaminergic denervation (positive DaT-SPECT group) while 52 patients did not (negative DaT-SPECT group) (Fig. 3A). Positive and negative group patients did not differ regarding age, sex or time since onset of symptoms (Table 3). Positive group patients presented higher bradykinesia and rigidity scores, but the two groups were not different regarding total tremor and RT MDS-UPDRS scores (Table 3). Next, we analysed the data collected from the motion sensor positioned in the lower back (Fig. 3B). The raw acceleration was filtered, a principal component analysis was performed, and the first component was used as the main axis of movement (Full details in the methods section). Given the oscillatory nature of tremor and in line with previous work (37,38), we used spectral analysis of the accelerometer data to compare the different groups (Fig 3B, bottom).

**Figure 3.**
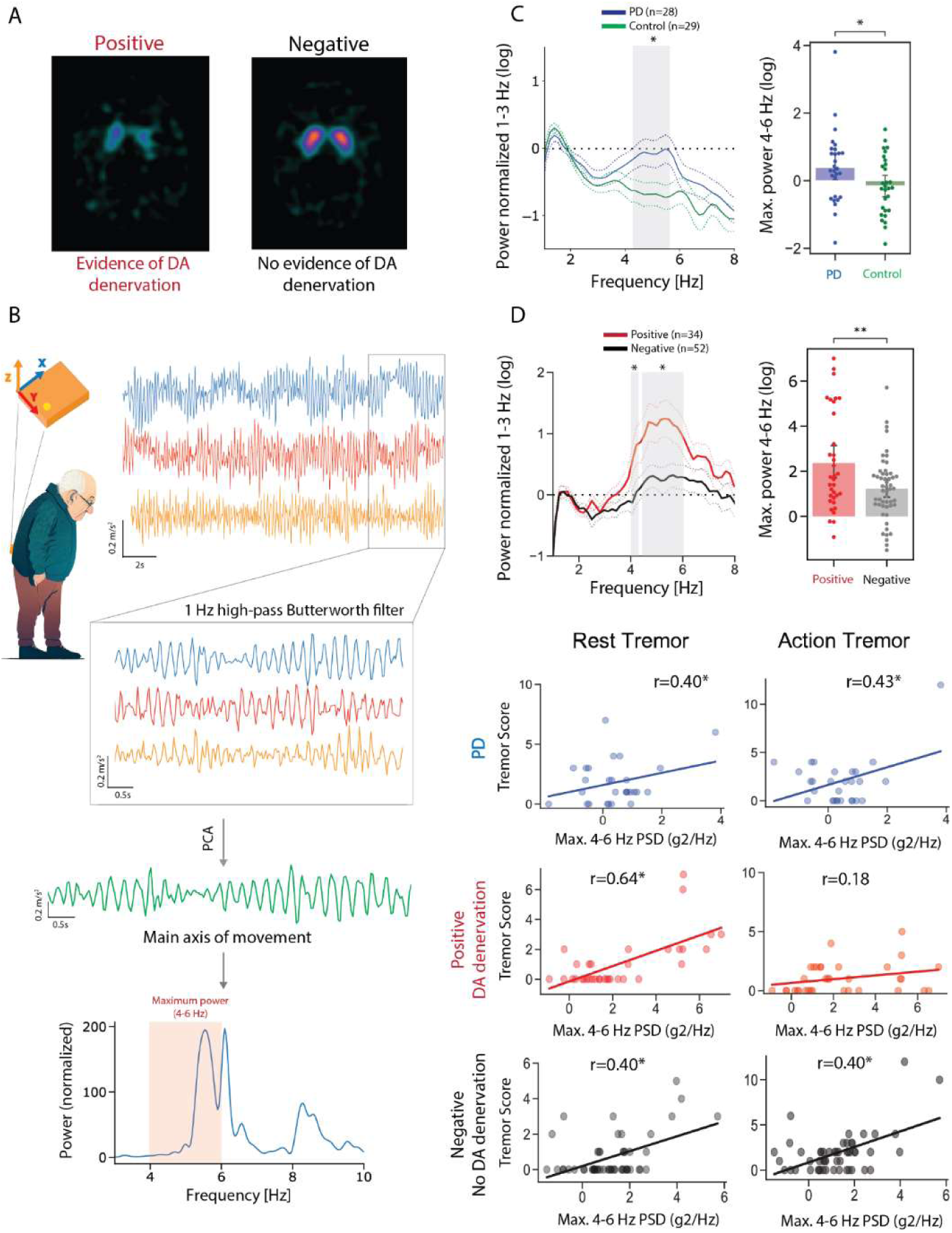
Oscillations assessed by inertial sensors are associated with higher Caudate Binding Potential. **(A)** Champalimaud Clinical Centre DaT-SPECT images were classified into positive (evidence of dopamine denervation) or negative (no evidence of dopamine denervation) based on standard clinical criteria **(B)** Inertial sensor placement and orientation in patients’ lower back. Example of triaxial acceleration, along with the schematic representation of the signal pre-processing pipeline and extraction of the maximum spectral power between 4-6 Hz, with an example of the Welch power spectral density (bottom). **(C**) Left, Welch power spectrum (normalized to the mean power between 1 and 3Hz) for the PD (blue) and Control (green) groups. Right, Comparison of the log of the maximum power spectrum **(D**) Left, Welch power spectrum (normalized to the mean power between 1 and 3Hz) for the Positive (red) and Negative (grey) groups. Right, Comparison of the log of maximum power spectral density in the 4-6 Hz band between the Positive and Negative groups. **(E)** log of maximum power spectral density (4-6 Hz) vs. Rest (left) and Action (right) tremor scores for PD, the Positive and Negative groups.

Considering the Welch power spectral density of standing accelerometry, an interaction was found between group and frequency in the PD/Control cohort (Repeated Measure Mixed-Effect Model, Group: *F*_1,55_=2.530, *P*=0.1174; Frequency: *F*_44,2420_=11.98, *P*<0.0001; Interaction: *F*_44,2420_=2.037, *P*<0.0001, Figure 3C) and also in the DaT-SPECT cohort (Repeated Measure Mixed-Effect Model: DaT-SPECT Group: *F*_1,84_=4.727, *P*=0.0325; Frequency: *F*_4400,3696_=12.84, *P*<0.0001; interaction: *F*_44,3696_=2.615, *P*<0.0001, Fig. 3D). To further explore the interaction, we performed post-hoc tests for each frequency bin in each group. We found that both the PD and the positive DaT-SPECT group significantly differed from the control and negative DaT-SPECT group, respectively, in almost all the bins in the 4-6 Hz band, the characteristic frequency band of RT in PD. Because tremor is not represented by a distinct resonance peak in the power spectral density(39), and due to the known distribution of frequency representation of RT in PD, we used the log of maximum power spectral density between 4 and 6 Hz as a metric to characterize oscillations within this frequency band. We found that this metric was significantly different between PD and Controls (Fig 3C, PD: 0.4±1.1, Controls: −0.2±0.9, *P*=0.037) and positive and negative patients (Fig 3D, 2.4±2.2 vs 1.2±1.4, *P*=0.01). After controlling for disease duration, this metric correlated with RT amplitude in both PD (*r*=0.40, *P*=0.04), positive (*r*=0.64, *P*<0.01) and negative (*r*=0.40, *P*<0.01) groups (Figure 3C). This was not tested in Controls as, per definition, no-tremor condition was present. As this metric was also found to be differentially associated with other motor symptoms (Supplementary Figure 3A), a linear regression model was performed where the 4-6 Hz maximum spectral power was used as a dependent variable and clinical symptoms as independent ones. When controlling for other motor symptoms, only RT remained associated with the 4-6Hz power positive DaT-SPECT: *P*<0.001, negative DaT-SPECT: *P*=0.023, PD: *P*<0.050 Supplementary Figures 3B-E).

These analyses demonstrate that in the context of DA denervation, a relatively specific 4-6 Hz oscillation is found, and it converges with the clinically assessed construct of RT.

### Integrity of caudate dopaminergic terminals is associated with the amplitude of rest oscillations

Given the correlation of our oscillation based metric with RT, we tested its association with the integrity of DA terminals. In positive patients, this metric was significantly correlated with CBP (*r*=0.41, *P*=0.02), but not with PBP (*r*=0.22, *P*=0.22, Figure 4A). This was not seen in negative patients (*r*=0.07, *P*=0.61; *r*=0.14, *P*=0.31, respectively, Fig 4A). Even when we focused on the small group of patients with a positive scan and no clinically defined RT (total RT = 0), the association between our metric and CBP was positive and of similar effect size, although non-significant (*r*=0.37, *P*=0.13 Supplementary Figure 4B). Interestingly although a positive correlation was also seen between MDS-UPDRS III-assessed RT and CBP, it had a lower effect size and was non-significant. (*r*=0.26, *P*=0.15, Supplementary Fig 4A). The lack of significance in this result may reflects both the lower N of this cohort and the limitations of the MDS-UPDRS III to assess the biological phenomenon. In corroboration, the well-known relationship between bradykinesia and PBP (*r*=-0.19, *P*=0.28, Supplementary Fig 4C) had a similar magnitude but was also not significant in this cohort. No relevant trends were found for PBP or negative patients (Supplementary Fig 4C-D).

**Figure 4.**
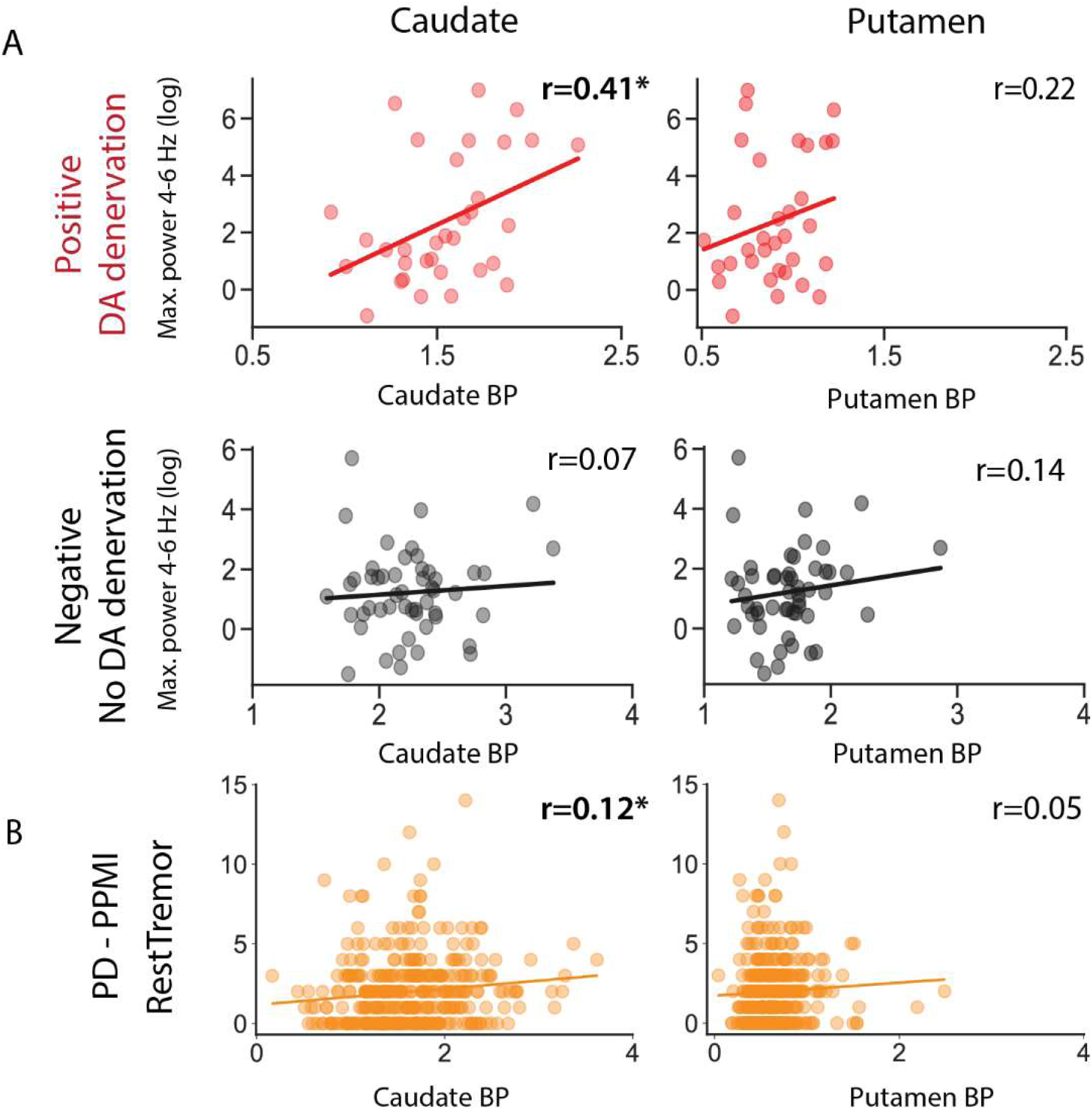
Oscillations assessed by inertial sensors are associated with higher Caudate Binding Potential. **(A)** log of maximum power spectral density (4-6 Hz) vs. CBP (left) and PBP (right) for the Positive and Negative groups. (B) MDS-UPDRS III assessed RT vs CBP (left) and PBP (right) for PPMI PD patients.

Power limitations can be reduced by taking advantage of the large PPMI dataset. In this dataset, RT amplitude was found to be positively associated with CBP (*r*=0.124, *P*=0.02) but not PBP (*r*=0.051, *P*=0.32, Fig 4B). A linear regression model found that even when adjusted for relevant clinical variables, RT amplitude was significantly associated with CBP (*P*=0.0108, Supplementary Table 3). However, this correlation is fueled by the presence of patients witthout RT, since the significant association between RT amplitude and CBP disappeared when we only considered patients with RT>0 (r=0.039, P=0.53).

Considering the lateralized nature of the basal ganglia motor control, we expanded our analysis to consider lateralized associations between RT and DA terminals integrity.

### A positive correlation between ipsilateral CBP and RT scores can be explained by higher CBP in patients with tremor

Multiple studies have tried to address the link between DA system and RT, with tremor-dominant forms being associated with more “benign” disease forms. Taking advantage of a recent systematic review on neuroimaging in PD(40), we performed a meta-analysis on studies where CBP was compared between patients presenting with a tremor-dominant (TD) phenotype vs. akinetic-rigid (AR) (Fig 5A) (22–27,41–44). For this analysis, the average CBP was extracted in each of the groups and the ratio was calculated. Ratios above 1 mean higher CBP in tremor-dominant patients. Nine out of 11 (one without data available) reported a higher CBP in tremor-dominant patients. Average ratios were 1.11±0.05 for contralateral CBP (9 studies, 47.3±22.7 patients per study), 1.09±0.06 for ipsilateral CBP (8 studies, 48.9±22.9 patients per study) and 1.09 for the only study with no side distinction (n=231). In our 2 datasets with DaT-SPECT data available, we separated patients, not by TD/AR phenotype but by the presence or absence of RT. In the PPMI cohort a rate of 1.11±0.02 was found and in the DaT-SPECT cohort a rate of 1.13±0.03.

**Figure 5.**
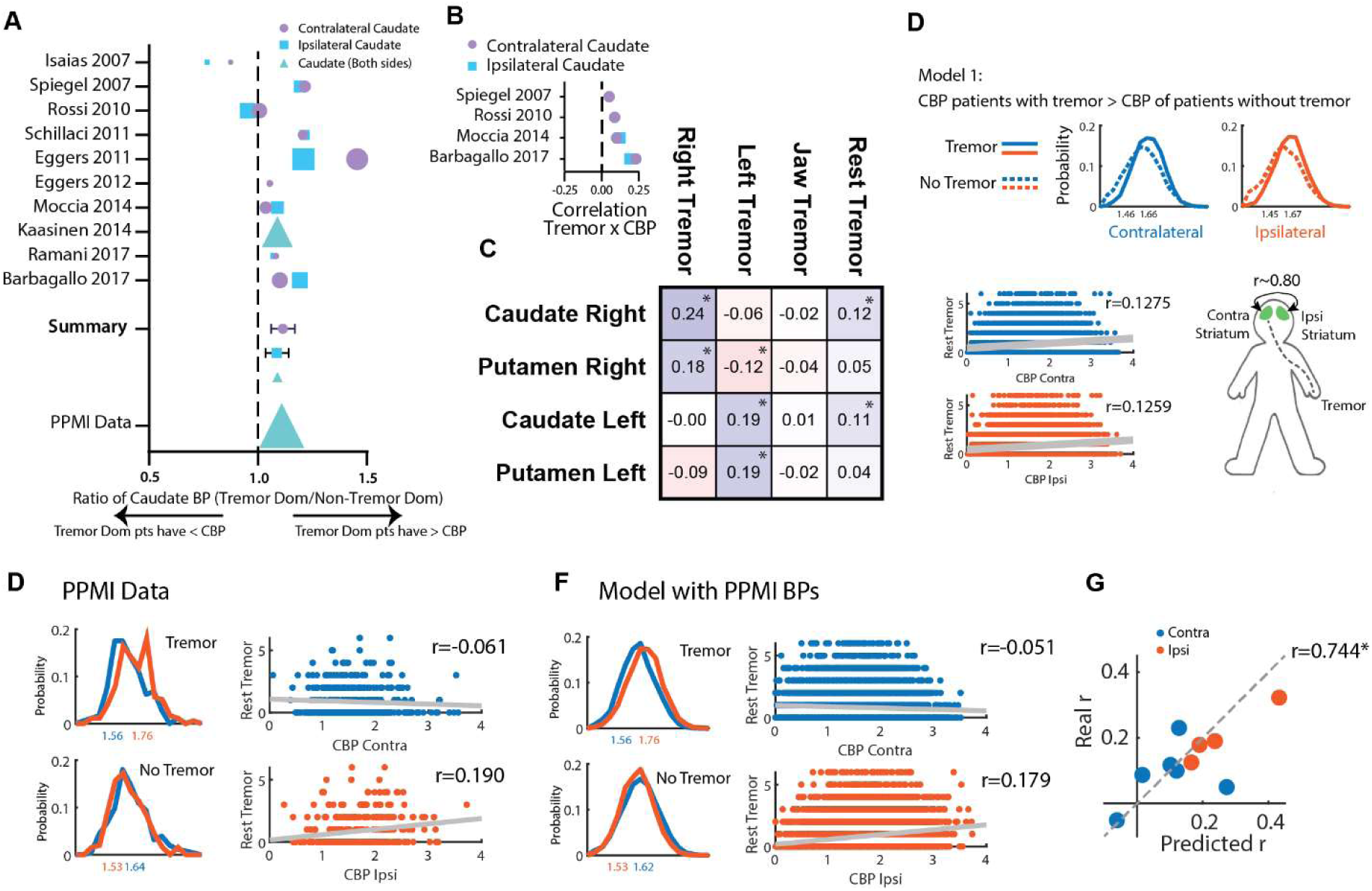
A higher CBP in patients with RT and asymmetry in caudate binding is sufficient to explain observations associating CBP with tremor amplitude. **(A)** Results from a meta-analysis on studies reporting CBP in different motor phenotype subgroups of PD patients (with Tremor/Tremor Dominant and without Tremor/No-Tremor Dominant). **(B)** Correlation coefficients between RT and CBP extracted from the literature. **(C)** Correlation matrix between RT related-symptoms (Right Tremor: Sum of Right Upper - 3.17a - and Lower Limb - 3.17c - RT scores; Left Tremor: Sum of Left Upper - 3.17b - and Lower Limb - 3.17d - RT scores; Jaw tremor: 3.17e; RT: Total sum of all scores) and Right and Left Caudate and Putamen binding potentials. **(D)** An *in silico* model was created with 3 assumptions: 1) Patients with tremor have a higher CBP than patients without tremor, 2) Tremor amplitude (i.e, values above 0) on the reference side is not associated with CBP, 3) Ipsi and contralateral CBP are highly correlated (∼0.80). Top: Simulated contralateral and ipsilateral CBP distributions for Tremor and No-Tremor groups, Bottom: Correlation of the simulated RT score and CBP. **(E)** Left: Distributions of ipsi and contralateral CBP in patients presenting tremor on the reference side and in patients presenting without tremor on the reference (PPMI data). Right: Correlation between tremor amplitude score and ipsi and contralateral CBP for the PPMI dataset. **(F)** Results from the model presented in D with distributions means defined based on the PPMI data described in G. Correlation results of a simulation of 10.000 subjects. **(G)** The same strategy used in F was applied to the data available and described in A. The scatter plot denotes the correlation of the coefficients predicted by our model with the coefficients reported in the original studies.

In our 2 cohorts, we also saw global results showing a positive association between oscillation power, tremor amplitude and CBP. Taking advantage of the same systematic review we collected data on studies correlating CBP with tremor amplitude(22,25,43,44). All report positive (even if non-significant) associations between tremor amplitude and CBP (Fig 5B).

In PD, symptoms frequently start on one side and the disease remains relatively asymmetric at least during the first years(45). If direct causality exists between caudate terminals integrity and RT severity, a lateralized finding is expected to emerge. In line with our general results, both right and left CBP were positively and significantly associated with total RT score (Fig 5C). This was not seen for the putamen. However, we were struck by the finding that this was driven by positive ipsilateral correlations. Right and left RT scores were positively associated, respectively, with right (*r*=0.24, *P*<0.001) and left (*r*=0.19, *P*<0.001) CBP (Fig. 5C). This result was unexpected and not aligned with current knowledge on basal ganglia motor control circuits where contralateral representation has been consistently described.

Having found that the presence of RT is related to higher CBP, we questioned if the ipsilateral correlations between RT and CBP could be simply the result of the combination of 1) a globally higher CBP in patients with any type of RT and 2) a high correlation between contralateral and ipsilateral CBP (as found in the PPMI cohort, *r*=0.779, *P*<0.001).

To achieve this, we created an *in silico* model with 3 main assumptions 1) Patients with RT have a higher CBP than patients without, 2) Tremor amplitude (i.e, RT values above 0) on the reference side is not associated with CBP, 3) Ipsi and contralateral CBP are highly correlated (∼0.80). With this model, we demonstrate these assumptions are enough for a positive ipsilateral correlation with tremor on the reference side to emerge (Figure 5D) but do not explain the absence of a contralateral correlation.

PD is known to be an asymmetric disease. If we restrict our analysis to patients selected by having any parkinsonian symptom on the reference side (100% of subjects have at least a unilateral disease) we are sure that in a large population, these symptoms will be less common contralaterally (<100% of subjects have a bilateral disease). If the tremor is present on the reference side, there is likely a nigro-striatal lesion contralaterally (therefore, a lower average contralateral, than ipsilateral PBP and CBP), but, at a population level, less severe DA depletion ipsilaterally. These, and other intuitions were supported by the data in the PPMI cohort (Supplementary Figure 5) and led us to the last assumption: 4) PD patients with RT on one side have a lower contralateral CBP, and PD patients without tremor on the reference side have a lower ipsilateral CBP. This assumption was evident when we analyzed the PPMI dataset (Figure 5E).

Using the left side as a reference and the mean and standard deviation of CBP in the PPMI dataset (Figure 5E) we simulated CBP values (Figure 5F) with all 4 assumptions. We found that the emerging correlations from our simulations were very similar to the ones originating from real data (real data: CBP contralateral, *r*=-0.061, ipsilateral, *r*=0.190; simulated: CBP contralateral, *r*=-0.051, ipsilateral, *r*=0.179). Details on further validation of this model can be found in Supplementary Figure 5.

Performing a meta-correlation, that was based on data available in the literature (Figure 5B), the PPMI dataset and our dataset, we found that this theoretical model based on CBP distributions in the tremor/tremor dominant or no-tremor/no-tremor dominant groups is highly efficient to predict the correlations between CBP and tremor amplitude (Figure 5G, *r*=0.744, *P*=0.0135).

## Discussion

RT remains one of the most puzzling symptoms in PD. Not only due to its relative disease-specificity but also due to the heterogeneity of responses to treatment(7,46). Current evidence has placed two main circuits in the center of tremor pathophysiology(16): The basal ganglia and cerebello-thalamo-cortical circuits with the thalamic VIM linking them. This model has been very effective in reconciling previous evidence and explaining why tremor produced by the cerebello-thalamic loop (the dimmer) is only seen in the presence of a dopaminergic dysfunction possibly in the pallidum (the switch). However, this model still does not reconcile major phenomenological observations: it doesn’t completely explain how levodopa replacement affects tremor amplitude (suggesting a dimmer and not only a trigger role for the DA system) or it also doesn’t clarify the multiple observations linking muscarinic receptors antagonism and tremor improvement.

We found that in PD a higher integrity of DA terminals in the caudate, but not the putamen, is related to the presence of RT. Although we disclose multiple pieces of evidence suggesting that the amplitude of oscillations (or RT severity) is positively related to the sparing of caudate terminals we believe detailed studies remain necessary. Even if our *in silico* model suggests that most of the association effect between these dimensions may be driven by a simple link between an RT phenotype and the integrity of caudate terminals, we need to keep in mind that tremor scales behave logarithmically(47). It is reasonable to consider that facing a clinical RT score of 0, variability in biological 4-6 Hz oscillations power is present (Suppl Figure 4B). This could be the necessary high-resolution to increase power in studying links between biological findings and strengthen current models. In fact, in the PPMI cohort, we found that caudate DA terminals relative sparing is related to the emergence of RT. We hypothesize that this could be related to the transition from a “clinically undetectable state” (RT=0) to a “clinically detectable state” (RT>0) aligned with a higher CBP phenotype. Longitudinal usage of IMUs could be relevant to assess this hypothesis.

Caudate is frequently considered the main non-motor (cognitive/associative) input structure to the basal ganglia, linked with goal-directed actions, learning, inhibitory control and other emotional roles(48,49). However, projections from the caudate to thalamic VIM have been described(50) and they seem to be of higher volume than those of putamen. These observations rely, not only on anatomical tracing evidence but direct neurophysiological manipulations(50–52). The caudate is also particularly well placed to link evidence of the cholinergic system involvement in tremor with current models. Anticholinergic agents were the first drugs available for symptomatic treatment of PD – their original use dates to the time of Charcot(53). Anticholinergic agents (trihexyphenidyl, benztropine, biperiden, ethopropazine, etc) help reduce all symptoms of PD, but they have found special favor in reducing the severity of tremor(46,54). Simultaneously, pro-cholinergic agents such as rivastigmine increase tremor amplitude(55). Intracaudate injection of muscarinic agonists induces tremor in rats, cats and monkeys(56–59) with a simultaneous increase in dopamine release(60,61), which is reverted by antimuscarinic agents. By parallelism, intracaudate injection of dopamine-depleting agents(32) also induces tremor, that is not increased with additional local administration of acetylcholine but is reverted by antimuscarinic agents. This suggests that this caudate-generated tremor involves a local cholinergic mechanism with a complex and precise fine-tuning between cholinergic and dopaminergic systems. Changes in oscillatory medium spiny neurons (MSN) activity propagates through basal ganglia circuits and possibly directly to VIM, but any interpretation of DA replacement effect is complicated by the influence of DA on tremor mechanisms generated by the pallidum(15) and thalamic nuclei(62).

Tremor emerges from an oscillatory network that involves the thalamus, motor cortex, basal ganglia and cerebellum. These are fast GABAergic and glutamatergic circuits that can causally contribute to oscillatory activity. We would argue that the role of caudate dopamine in this circuit is different: DA is a neuromodulator that facilitates the oscillatory system to enter specific states. These states are promoted by an interaction between multiple modulatory systems (caudate Ach/DA, pallidal DA, noradrenaline(28) and serotonin(29,30)). These slower properties would be hard to capture using a second-by-second functional magnetic resonance imaging (fMRI) analysis paired with the tremor signal but could emerge when slower timescales or tremor-independent designs are used. A fMRI study has found that during a force task, tremor-dominant PD patients present a higher activation of the contralateral caudate nucleus(63) while activity in the caudate nucleus as assessed by ^18^F-FDG positron emission tomography (PET) was related to tremor and modified when VIM DBS On/Off were compared(64). Again, this evidence emerges from manipulations that are non-causal, but support a role for caudate activity (modulated by DA) on tremor pathophysiology. The activity of the caudate (or striatum) has not been included in the strongest fMRI studies using a causal dynamic model approach(62,65–67). We believe that integrating this node in future analysis could provide valuable insights into its role in RT pathophysiology.

DA has been proposed to have a role in reducing RT by directly facilitating inhibitory mechanisms in thalamic VIM(62). The thalamic VIM receives dopaminergic projections from both A8 (RRF) and ventral A9 substantia nigra pars compacta (SNc) nuclei(68) regions known to project also to anterior caudate nuclei(68–70) and putamen. These observations place the relationship of DA with RT in a multidimensional space where it is very likely that a combination of the topography of DA neuron loss and the local integrity of DA terminals are the key elements for RT to emerge and to explain its’ therapeutic response. The low spatial resolution of SPECT provides a limited ability to detect these patterns. Multimodal imaging with ^18^F-FDOPA PET(71) and quantitative MRI(72) may be the necessary tools to assess these symptoms and in vivo neuropathological correlates in more detail. At last, as this evidence is correlative, we should never exclude the hypothesis that the integrity of dopaminergic terminals in the caudate is a proxy of causal DA dysfunction in other nodes of the circuit.

Rating scales are a strong and useful tool for clinical practice – they capture disease severity and provide a tool for disease assessment across multiple centres. However, assessment bias may exist and MDS-UPDRS change scores contain an important amount of error variance^62^. Additionally, biological systems do not necessarily behave linearly, as is the case of the logarithmic relationship identified between tremor rating scales and tremor amplitude(73). This may relate to the difference in the coefficient of determination (R^2^) we found when clinically assessed tremor is related to CBP (≈0.01 – Fig 4A – to 0.07 – Supplementary Fig 4) when compared with an IMU-based tremor assessment (≈0.17 – Fig 4B), suggesting that the use of unbiased assessment metrics can provide a refinement that enables a better understanding of tremor pathophysiology (and potentially other motor symptoms).

Our *in silico* model was able to replicate the coefficients of correlation between ipsi or contralateral CBP and RT in the PPMI dataset, in data we collected and 4 other published studies (22,25,43,44). This is an important reminder of the correlative nature of this type of analysis. Although it is easier to be motivated to seek trivial explanations for a biologically implausible correlation like the one described, more biologically reasonable correlations can also have a spurious origin in bilateral systems. Our results show that positive ipsilateral associations between CBP and tremor amplitude are a trivial result driven by the asymmetric nature of PD and higher total CBP in patients with tremor.

Limitations in our study should be acknowledged. Our DaT-SPECT cohort and the PPMI cohort are biased for early disease stages. Our findings regarding tremor mechanisms may not be generalizable for later stages (aligned with the contrast we observed with the clinical PD populations). Besides the high affinity for DaT, ioflupane is known to have a tenfold lower affinity for serotonin transporter (SERT)(74). We cannot exclude that part of the effect we found is driven by some lack of specificity and binding to SERT, however, tremor in PD has been linked to a reduction in basal ganglia SERT binding (75), and not an increase. Thus, our results are very unlikely to be driven by SERT binding.

Specific vulnerability of dopaminergic neurons has been a major focus in PD research(76,77). Evidence on specific functional roles(78,79) and anatomical projections(80) of SNc neurons is mounting, and their function and vulnerability may be linked to specific genetic identity(81). Tremor has been linked with PD forms with a better prognosis. Here, we present data supporting that relative caudate DA integrity is linked with this specific symptom and hypothesize that this may emerge due to a lower vulnerability of caudate projecting dopaminergic neurons to progressive degeneration. Individual genetic heterogeneity interacting with the vulnerability associated with the specific genetic signature of a neuron may lead to different subpopulation losses and therefore different phenotypes(82). Within this model, tremor may emerge as a byproduct of specific circuits integrity and general neuronal resilience to death. A better understanding of the tremor circuit and the refinement of the current model of PD RT can provide important insights into PD pathophysiology.

## Methods

Our study examined both male and female subjects. Findings are reported for both sexes.

### PPMI cohort

#### Subjects and Clinical Assessment

We accessed the Parkinson’s Progression Markers Initiative (PPMI) database (http://www.ppmi-info.org/) and selected all patients that presented our inclusion criteria: data available at visit 6 (24 months) regarding the revised Movement Disorder Society Unified Parkinson’s Disease Rating Scale (MDS-UPDRS)(35) part III RT score with an OFF assessment. A total of 432 patients were included. Clinical data was extracted from visit 6 (year 2), visit 4 (year 1) and baseline (or screening visit if adequate, year 0).

Motor ratings were obtained using the MDS-UPDRS. Part III of the MDS-UPDRS was used to score patients’ rigidity (total score on item 3.3), bradykinesia (total score on items 3.4, 3.5, 3.6, 3.7, 3.8, 3.9 and 3.14), action tremor (total score on items 3.15 and 3.16) and RT (total score on item 3.17). Patients were classified as RT present or absent based on the total score on item 3.17 (>0 or 0). For all analyses, OFF state evaluations were used.

#### ^123^I-FP-CIT SPECT protocol and analysis

DaT imaging was obtained using single-photon emission computed tomography (SPECT) after ^123^I-FP-CIT intravenous injection. Binding ratios were extracted from those already processed centrally. Briefly, SPECT raw projection data was imported to a HERMES (Hermes Medical Solutions, Skeppsbron 44, 111 30 Stockholm, Sweden) system for reconstruction using the ordered subsets expectation maximization (OSEM) algorithm. This was done for all imaging centres to ensure the consistency of the reconstructions. Reconstruction was done without any filtering applied. The OSEM reconstructed files were then transferred to the PMOD (PMOD Technologies, Zurich, Switzerland) for subsequent processing. Attenuation correction ellipses were drawn on the images and a Chang attenuation correction was applied to images utilizing a site-specific μ that was empirically derived from phantom data acquired during site initiation for the trial. Once attenuation correction was completed a standard Gaussian 3D 6.0 mm filter was applied. These files were then normalized to standard Montreal Neurologic Institute (MNI) space so that all scans were in the same anatomical alignment. Next, the transaxial slice with the highest striatal uptake was identified and the 8 hottest striatal slices around it were averaged to generate a single-slice image. Regions of interest (ROI) were then placed on the left and right caudate, the left and right putamen, and the occipital cortex (reference region). Mean counts per voxel for each region were extracted and used to calculate binding potentials (BP) for each of the 4 striatal regions: left and right caudate binding potential (CBP) and putamen binding potential (PBP). The BP was calculated as 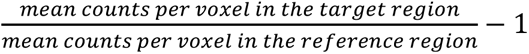.

We extracted data from DaT SPECTs performed at visit 0/screening, visit 4 (1 year after) and visit 6 (2 years after).

### Clinical cohorts

#### Hospital Egas Moniz Subjects and Clinical Assessment

A total of 57 patients were consecutively recruited from the General Neurology and Movement Disorders outpatient clinic at Hospital Egas Moniz in Lisbon, in the period from January 2019 to June 2019. We consecutively recruited patients with a diagnosis of clinically probable Parkinson’s disease according to the MDS clinical diagnosis criteria(83). Patients were recruited independently of disease duration and/or characteristics. We excluded patients incapable of walking without aid.

Healthy controls (HC) were consecutively recruited from non-consanguineous family members or caregivers attending the outpatient clinic. Controls (*n*=29) were included if they had no known diagnosis of a neurological disorder (excluding headache) and, on a neurological evaluation, did not report any symptom or presented any sign suggestive of a motor disease. Controls and PD patients were age-matched (PD/Control cohort).

Participants were clinically evaluated by three of the investigators (MM, RB, BM), who collected demographic and clinical data and administered the MDS-UPDRS part III.

#### Champalimaud Clinical Centre Subjects and Clinical Assessment

A total of 90 patients were consecutively recruited from the Nuclear Medicine clinical service at the Champalimaud Clinical Centre in the period from February 2019 to July 2023. Four (4) patients were excluded due to loss/corruption of kinematic or clinical data. A total of 86 patients were considered for the present study. We recruited 1) patients referred to a ^123^I-FP-CIT SPECT for the differential diagnosis of a movement disorder and 2) were at least 18 years old. We excluded patients referred for differential diagnosis of dementia. Recruitment was interrupted between 2020 and 2022 due to the COVID-19 pandemic. Patients were excluded if the met any of the exclusion criteria: 1) dementia, painful arthritis, peripheral neuropathy or any disorder that may influence walking, 2) any relevant unstable medical condition per investigator judgement, 3) used a walking aid, 4) were pregnant, and 5) had any contraindication for ioflupane.

Participants were clinically evaluated by two of the investigators (MM, JAS), who collected demographic and clinical data and administered the MDS-UPDRS, parts II and III. Motor ratings on both clinical cohorts were computed from the MDS-UPDRS part III as described for the PPMI cohort.

#### Inertial sensor assessment and analysis

Patients’ movement was assessed with a lower back-mounted Xsens MTw wireless inertial sensor (Movella, Las Vegas, USA). The sensor contains a three degrees of freedom accelerometer, gyroscope and magnetometer. Inertial data was captured at a resolution of 100 Hz at the Egas Moniz cohort and at a resolution of 40 Hz at the Champalimaud cohort while patients performed a standing task. In the Egas Moniz cohort, the task consisted of standing still for 30 seconds. In the Champalimaud cohort, the task consisted initially of standing still for 30 seconds (45 patients) and later, it was updated to 60 seconds, to capture a longer period of postural stance (41 patients). During the task, raw inertial data were logged and synchronised by an MTw Awinda (Movella, Las Vegas, USA) station and saved to a laptop.

Using rotation matrices, raw accelerometer data were first rotated so that the Z-axis pointed downwards (positive Z-axis in the direction of gravity). To avoid signal rotation artifacts, one hundred (100) data points were trimmed from both ends of the rotated signal. Then, all three axes were low-pass filtered using a 5^th^-order Butterworth filter with a 1 Hz frequency cut-off to remove the gravitational acceleration component. Oscillations resulting from tremor may occur in a combination of all degrees of freedom. To capture oscillations irrespective of their direction, we performed a principal component analysis (PCA) using the three acceleration axes, from which the first principal component yields the axis of greatest variance, which provides an approximation of the axis that captures most oscillations(84,85). An estimate of the power spectral density of the first principal component was then computed using the Welch method(86) normalized to the low frequency component of the signal (power between 1-3 Hz), from which we then extracted the natural logarithm of the maximum power between 4-6 Hz. This band is described as the frequency band where Parkinsonian RT typically occurs(87). Due to its skewed distribution, this metric was logarithmically transformed to compare between groups. All accelerometer pre-processing and analysis were performed in Python version 3.8.15.

### DaT SPECT protocol and analysis

All subjects in the Champalimaud Cohort were pre-treated with iodine to reduce the potential thyroid irradiation and then intravenously injected with ^123^I-FP-CIT. SPECT image acquisition was performed in a gamma camera (Philips BrightView) approximately 2 hours after ^123^I-FP-CIT administration. Images were classified into positive (evidence of dopamine denervation) or negative (no evidence of dopamine denervation) based on an evaluation performed by experienced nuclear medicine physicians. Furthermore, an automated quantification of regional binding potentials was conducted using validated software(88). In short, first, the brain DaT-SPECT volume is registered to a template image, and then a reference region with nonspecific uptake containing the entire cerebrum except the striatum and neighbour regions is defined. This method (instead of using an occipital or cerebellar reference) is less sensitive to noise or artifacts(89,90). Finally, two ROI on each brain hemisphere – one over the caudate and another over the putamen (Supplementary Figure 4A) – are automatically defined. Left and right caudate and putamen binding potentials (CBP and PBP) were computed independently for each hemisphere in a standard fashion as in the PPMI dataset.

### Literature review and analysis

We took advantage of the recent publication of a systematic review(40) to identify studies comparing neuroimaging differences between PD motor subtypes. We identified all studies using DaT-SPECT imaging and, for each study, we extracted data on the CBP in tremor-dominant groups and non-tremor-dominant groups. When more than 2 control groups were described, data from the Akinetic-rigid group was selected. When available we collected independent data from contralateral and ipsilateral caudate (to most severe symptoms). If not detailed, average values were extracted. For each study, the caudate binding ratio (CBR), computed between the mean CBP of the tremor group and the mean CBP of the no-tremor group was calculated. If CBR is higher in tremor groups, the value will be above 1, if lower, the value will be lower to 1. Results were averaged across studies with a weighting factor of 1 for each study. When a study presented the correlation coefficient between CBR and the RT subscore, this was also extracted.

### Statistics

Descriptive statistics were presented as means and standard deviations. For most longitudinal analyses where time was a variable, repeated measures mixed effect model was used with a second variable accounted for (tremor Group). These models considered within-subject correlation and between-subject variability and can account for missing data. Multiple comparison correction was performed with the Šídák’s or the Holm-Šídák’s multiple comparisons test as appropriate. When two groups with continuous variables were compared either paired or unpaired t-test was used accordingly. Welch correction was performed when group variances were unequal. For categorical variables, the Chi-square test was used. When a reference value was known, a one-sample t-test was used. Linear and logistic regressions were built based on *a priori* hypothesis on possible confounders and relevant variables. Correlations were performed using the Pearson *r* except when non-normality of the distribution was documented (in this context, Spearman’s rho was used). For the comparison of the power spectrum of the clinical cohorts, a two-way mixed design ANOVA was used and multiple comparisons between groups were done using Fischer’s least significant difference. The significance level was set at 0.05.

### Study approval

The study was approved by the Centro Hospitalar de Lisboa Ocidental Ethics Committee and by the Champalimaud Foundation Ethics Committee. Written informed consent was obtained from all participants before any study procedure.

## Data availability

The PPMI data is part of an open database. Access to PPMI data can be requested at https://www.ppmi-info.org/access-data-specimens/download-data. The data from the Champalimaud Clinical Centre cohort will be made available upon reasonable request.

Scripts of the Matlab and Python codes used for data analysis can be found at https://github.com/jalvesdasilva/Caudate_PD_Tremor

## Author contributions

MM and JAS designed the research studies with additional input from PF, DC, FO and AJOM. MM, PF, RB and BM collected the data from the Hospital Egas Moniz cohort, MM, PF and JAS collected the data from the Champalimaud cohort. MM analyzed the PPMI data with additional input from JAS. PF analyzed the 2 clinical cohorts data with input from MM and JAS. FO developed the tools for analysis of the SPECT results, with input from DC. MM developed the *in silico* model with input from JAS and PF. MM, PF and JAS wrote the first draft of the manuscript that was critically reviewed by all authors. MM and PF contributed equally. The author order was chosen based on the contributions to all elements of the study.

## Acknowledgements

Data used in the preparation of this article were obtained on May 13, 2023, from the Parkinson’s Progression Markers Initiative (PPMI) database (www.ppmi-info.org/access-data-specimens/download-data), RRID:SCR 006431. For up-to-date information on the study, visit www.ppmi-info.org.

PPMI – a public-private partnership – is funded by the Michael J. Fox Foundation for Parkinson’s Research and funding partners, including including 4D Pharma, AbbVie, AcureX, Allergan, Amathus Therapeutics, Aligning Science Across Parkinson’s, AskBio, Avid Radiopharmaceuticals, BIAL, Biogen, Biohaven, BioLegend, BlueRock Therapeutics, Bristol-Myers Squibb, Calico Labs, Celgene, Cerevel Therapeutics, Coave Therapeutics, DaCapo Brainscience, Denali, Edmond J. Safra Foundation, Eli Lilly, Gain Therapeutics, GE HealthCare, Genentech, GSK, Golub Capital, Handl Therapeutics, Insitro, Janssen Neuroscience, Lundbeck, Merck, Meso Scale Discovery, Mission Therapeutics, Neurocrine Biosciences, Pfizer, Piramal, Prevail Therapeutics, Roche, Sanofi, Servier, Sun Pharma Advanced Research Company, Takeda, Teva, UCB, Vanqua Bio, Verily, Voyager Therapeutics, the Weston Family Foundation and Yumanity Therapeutics.

MM, AJOM and JAS were supported by grant EurDyscover – EJPRD19-135, funded by FCT/MCTES through the European Joint Programme for Rare Disease (EJPRD/0001/2019). MM is supported by the Michael J. Fox Foundation (MJFF-023180). JAS was supported by the Portuguese Foundation for Technology and Science (FCT) CEEC grant (2020.03118.CEECIND). PF was supported by the Portuguese Foundation for Science and Technology (FCT) through a PhD scholarship (2023.03390.BD) and by the European Union’s Horizon 2020 Research and Innovation Programme through grant H2020-SC1-DTH-2019-875358 (FAITH project)

We thank Prof. Raúl Rato from Uninova (Faculty of Science and Technology, Universidade Nova de Lisboa, Lisbon) for an insightful discussion on spectral analysis of accelerometry data.

## Conflict of interest

MM has received in the past 3 years, payment, honoraria or other support from Bial, Pharmacademy, Evidenze and AbbVie.

A.J.O.-M. was a national coordinator for Portugal of a non-interventional study (EDMS-ERI-143085581, 4.0) to characterize a treatment-resistant depression cohort in Europe, sponsored by Janssen-Cilag, Ltd. (2019–2020), a trial of psilocybin therapy for treatment-resistant depression, sponsored by Compass Pathways, Ltd. (EudraCT number 2017-003288-36), and a trial of esketamine for treatment-resistant depression, sponsored by Janssen-Cilag, Ltd. (EudraCT NUMBER: 2019-002992-33). He is a recipient of a grant from Schuhfried GmBH for norming and validation of cognitive tests. In the past 3 years, he received payment, honoraria or other support from Angelini, Janssen, MSD, Neurolite AG, and the European Monitoring Centre for Drugs and Drug Addiction. He is vice-president of the Portuguese Society for Psychiatry and Mental Health and Head of the Psychiatry Working Group for the National Board of Medical Examination (GPNA) at the Portuguese Medical Association and Portuguese Ministry of Health. None of the aforementioned agencies had a role in the preparation, review, or approval of the manuscript or in the decision to submit the manuscript for publication.

Other authors report no conflict of interest.

Marcelo D. Mendonça and Pedro Ferreira contributed equally to this work Marcelo D. Mendonça and Joaquim Alves da Silva should be listed as corresponding authors.

