## Supplemental material for "Integrity of dopaminergic terminals in the caudate nucleus is relevant for rest tremor in Parkinson’s disease"

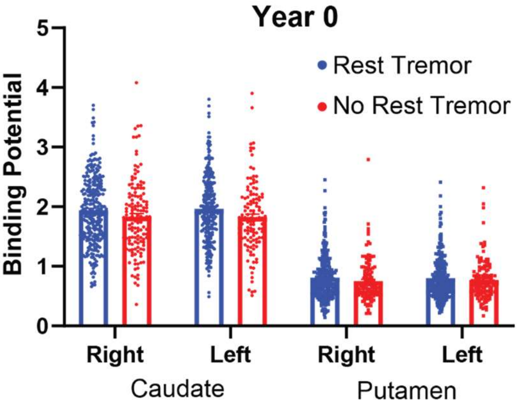


| Source of Variation | P value |
|---|---|
| Side | 0.8345 |
| **Brain Region** | **<0.0001** |
| **Presence of Tremor** | **0.0096** |
| Side x Brain Region | 0.9543 |
| Side x Presence of Tremor | 0.9452 |
| **Brain Region x Presence of Tremor** | **0.0079** |
| Side x Brain Region x Presence of Tremor | 0.298 |

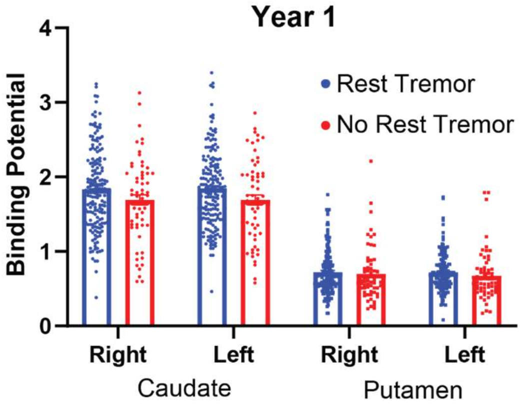


| Source of Variation | P value |
|---|---|
| Side | 0.884 |
| **Brain Region** | **<0.0001** |
| **Presence of Tremor** | **0.024** |
| Side x Brain Region | 0.625 |
| Side x Presence of Tremor | 0.8643 |
| **Brain Region x Presence of Tremor** | **0.004** |
| Side x Brain Region x Presence of Tremor | 0.8798 |

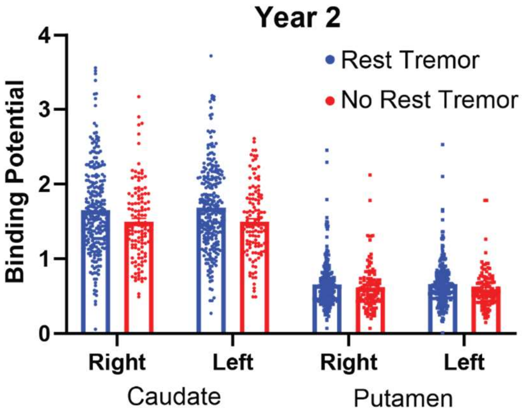


| Source of Variation | P value |
|---|---|
| Side | 0.9397 |
| **Brain Region** | **<0.0001** |
| **Presence of Tremor** | **0.0003** |
| Side x Brain Region | 0.3291 |
| Side x Presence of Tremor | 0.6219 |
| **Brain Region x Presence of Tremor** | **<0.0001** |
| Side x Brain Region x Presence of Tremor | 0.9619 |

**Supplementary Figure 1** – Three-way ANOVA analysis of DaT binding, performed at each timepoint. Groups were defined based on clinical classification at Year 2 (With RT or without RT). We found that there is consistently an effect of brain region (Caudate / Putamen), a group effect (Tremor / No-Tremor) and an interaction between them. This supports that the difference in BP observed in caudate is not present in putamen, irrespective of side.

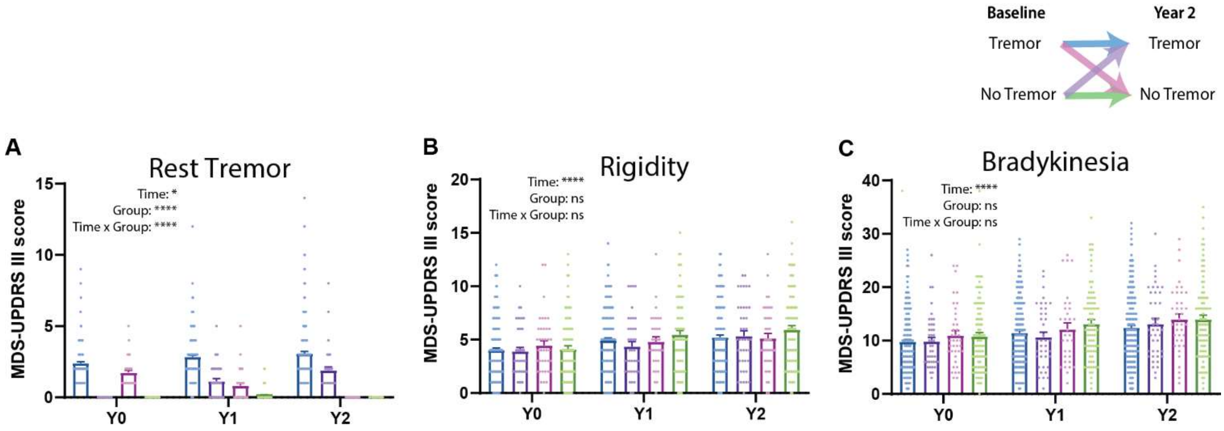


**Supplementary Figure 2 – A)** RT scores (MDS-UPDRS III 3.17 score). Time: *P*=0.0469, Group: *P*<0.0001, Time × Group: *P*<0.0001. Post-hoc (Šídák's multiple comparisons test), **B)** Rigidity scores (MDS-UPDRS III 3.3 score). Time: *P*<0.0001, Group: *P*=0.5771, Time × Group: *P*=0.1704. **C)** Bradykinesia scores (MDS-UPDRS III 3.4, 3.5, 3.6, 3.7, 3.8, 3.9 and 3.14 score). Time: *P*<0.0001, Group: *P*=0.1641, Time × Group: p=0.3139.

| | OR | 95% CI | *P*-value |
|---|---|---|---|
| Intercept | 0.282 | 0.116 – 6.224 | 0.425 |
| **Caudate Y2** | **7.235** | **1.592 – 39.060** | **0.014** |
| Putamen Y2 | 0.119 | 0.006 – 1.626 | 0.128 |
| Age | 0.980 | 0.940 – 1.021 | 0.329 |
| Bradykinesia Y2 | 1.007 | 0.931 – 1.088 | 0.863 |
| Rigidity Y2 | 0.979 | 0.8483 – 1.127 | 0.767 |

**Supplementary Table 1** – Logistic regression taking as dependent variable current presence of RT. Hosmer-Lemeshow test: *P*=0.342. AUC of the model: 0.654 (0.545–0.763; *P*=0.008).

| | Beta | 95% CI | *P*-value |
|---|---|---|---|
| Intercept | -0.013 | -1.776 – 1.750 | 0.988 |
| **Caudate Y2** | **0.864** | **0.174 – 1.555** | **0.014** |
| Putamen Y2 | -0.806 | -2.172 – 0.560 | 0.247 |
| Age | 0.013 | -0.010 – 0.036 | 0.259 |
| Bradykinesia Y2 | 0.011 | -0.032 – 0.054 | 0.616 |
| Rigidity Y2 | 0.040 | -0.044 – 0.124 | 0.355 |

**Supplementary Table 2** – Linear Regression taking as dependent variable the RT amplitude (as assessed by the MDS-UPDRS part III).

| | Beta | SE | *P*-value |
|---|---|---|---|
| Intercept | 2.050 | 0.180 | <0.001 |
| **Rest Tremor** | **0.033** | **0.013** | **0.011** |
| Rigidity | -0.008 | 0.010 | 0.409 |
| **Bradykinesia** | **-0.016** | **0.005** | **0.001** |
| Action Tremor | 0.004 | 0.015 | 0.809 |
| Age | -0.004 | 0.002 | 0.141 |

**Supplementary Table 3** – Linear Regression taking as dependent variable CBP

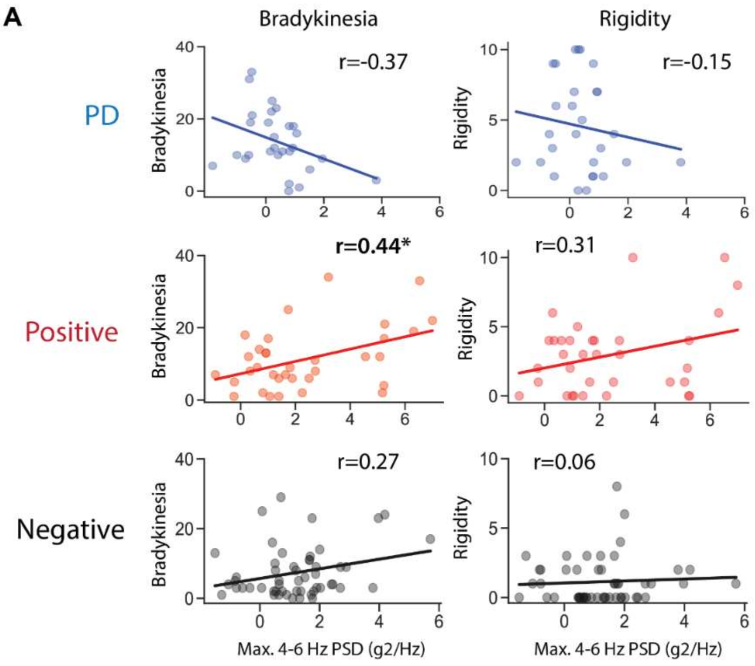


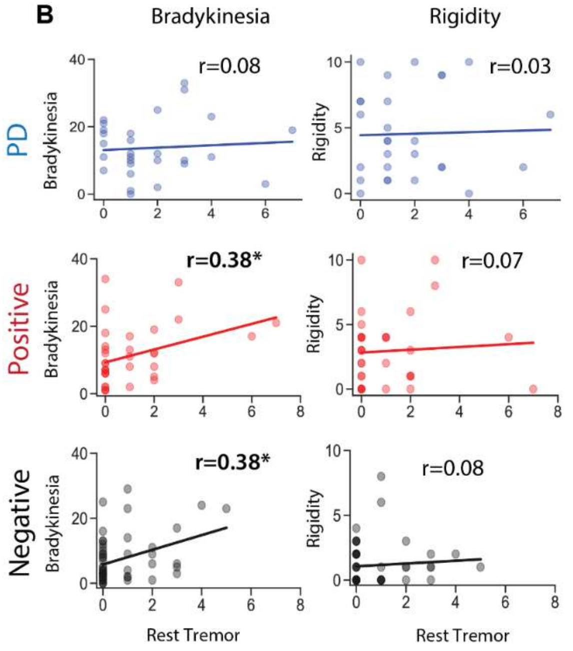


**C**

Association with Max. 4-6 Hz PSD (g2/Hz) — PD

| Variable | Estimate | 95% CI | p-values |
|---|---|---|---|
| Intercept | 0.7186 | -0.110 to 1.548 | 0.086 |
| **Rest Tremor** | **0.2367** | **8.1e-5 to 0.473** | **0.0499** |
| Bradykinesia | -0.0538 | -0.116 to 0.009 | 0.089 |
| Rigidity | 0.0416 | -0.109 to 0.192 | 0.574 |
| Disease duration | -0.0036 | -0.012 to 0.005 | 0.389 |

**D**

Association with Max. 4-6 Hz PSD (g2/Hz) — Positive

| Variable | Estimate | 95% CI | p-values |
|---|---|---|---|
| Intercept | 0.8744 | -0.076 to 1.825 | 0.070 |
| **Rest Tremor** | **0.9630** | **0.522 to 1.404** | **<0.001** |
| Bradykinesia | 0.0141 | -0.097 to 0.125 | 0.797 |
| Rigidity | 0.1874 | -0.130 to 0.505 | 0.237 |
| Disease duration | -0.0056 | -0.016 to 0.005 | 0.283 |

**E**

Association with Max. 4-6 Hz PSD (g2/Hz) — Negative

| Variable | Estimate | 95% CI | p-values |
|---|---|---|---|
| Intercept | 0.4811 | -0.115 to 1.077 | 0.111 |
| **Rest Tremor** | **0.4013** | **0.057 to 0.746** | **0.023** |
| Bradykinesia | 0.0243 | -0.036 to 0.085 | 0.420 |
| Rigidity | -0.0430 | -0.290 to 0.204 | 0.727 |
| Disease duration | 0.0076 | -0.000 to 0.015 | 0.055 |

**Supplementary Figure 3 – A)** There was a significant association between the 4-6 Hz maximum power and bradykinesia (*r*=0.44, *P*=0.01), but not with rigidity (*r*=0.31, *P*=0.07) for patients with a positive DaT-SPECT. **B)** Bradykinesia was significantly correlated with RT in both positive and negative groups (positive: *r*=0.38, *P*<0.05, negative: *r*=0.38, *P*<0.05). Conversely, no associations were found between rigidity and RT in either group (positive: *r*=0.07, *P*=0.71, negative: *r*=0.08, *P*=0.58). This was not seen for patients in the PD group (a more heterogeneous group). **C)**, **D)** and **E)** Linear regression models for the 3 groups (patients with and patients without evidence of dopamine denervation and PD patients) with the maximum power in the 4-6 Hz band as the dependent variable, and RT, bradykinesia and rigidity as predictors, found that, after adjustment to confounders only resting tremor was positively associated with maximum 4-6 Hz power.

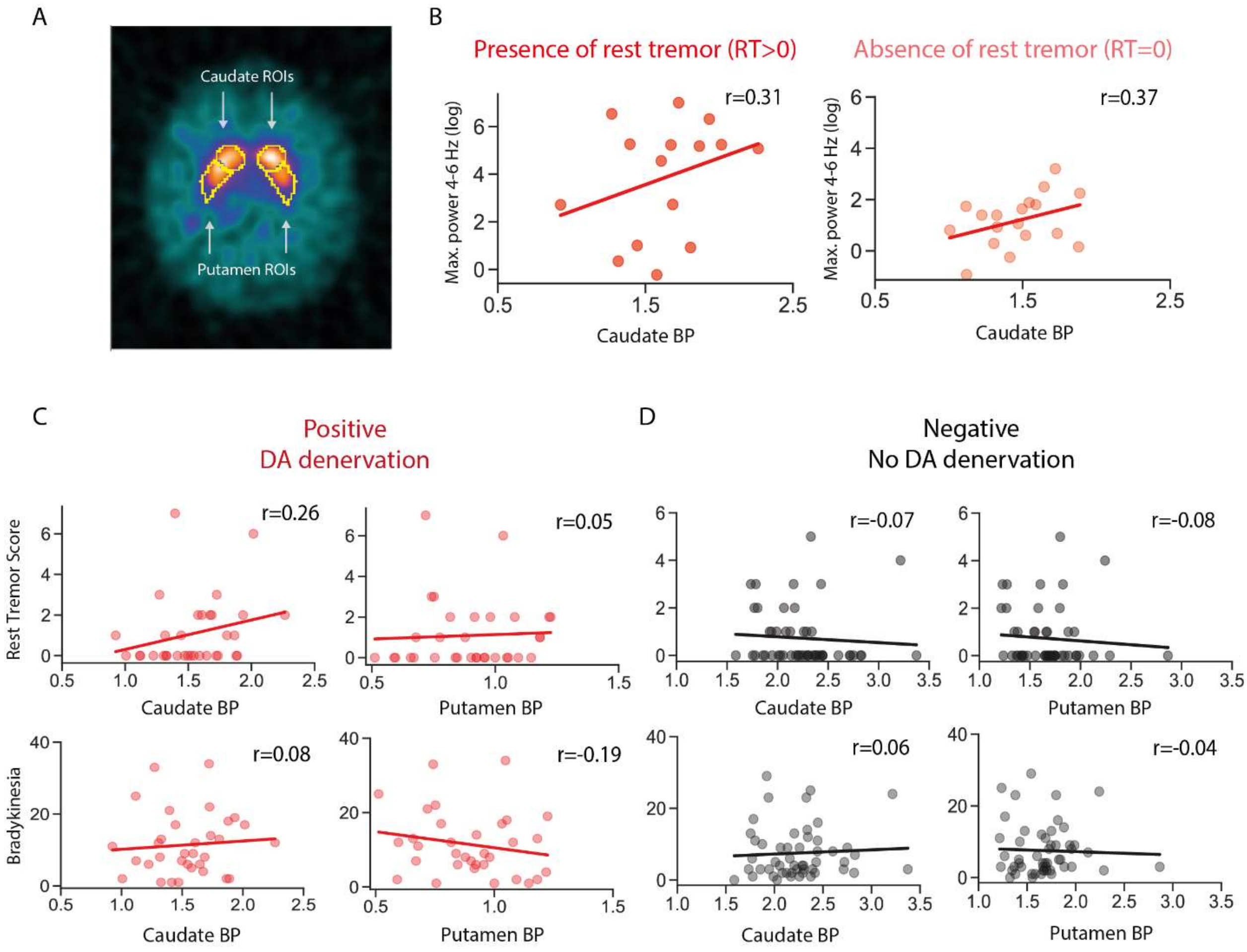


**Supplementary Figure 4 – A)** Representation of the ROIs corresponding to the caudate and putamen. **B)** A positive association between higher inertial oscillations and Caudate binding potential was seen when looking separately at patients with RT (*r*=0.31, *P*=0.26) and without RT (*r*=0.37, *P*=0.13). **C)** and **D)** Association between CBP (left) and PBP (right) and RT (top) and Bradykinesia (bottom) in the positive (DA denervation) and negative (no DA denervation) groups.

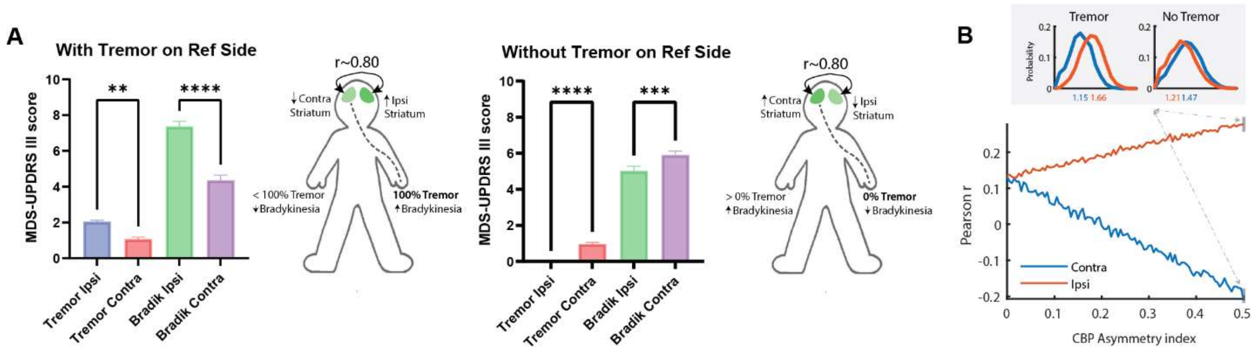

**Supplementary Figure 5 – A)** Data of PPMI patients at Y2 was divided into 2 groups: 1 group presenting tremor on a reference side (left side of the body) and another group without tremor in this reference side. If tremor is present on the reference side, it is very likely that there is a nigro-striatal lesion contralaterally (therefore, a lower contralateral PBP and CBP). Given that PD can be asymmetric and affect only one side in earlier phases, by definition, if tremor is present on one side, the probability of symptoms being present contralaterally is lower (therefore, a higher ipsilateral CBP). This intuition is supported by the PPMI data (left). Using these criteria, it's clear that patients with tremor on the reference side have more ipsilateral and less contralateral motor symptoms. By contrast, if tremor is not present in the reference side, in a population with two years of progression since diagnosis, it is very likely that contralateral symptoms are present. Aligned with contralateral symptoms we would expect a higher ipsilateral DA denervation. PPMI data also supports this intuition (right). Patients with no tremor on the reference side have more severe contralateral symptoms. We are also certain that all patients with tremor in the reference side correspond to a tremor population (thus, with a higher average CBP). Moreover, this population is, enriched in patients without tremor (therefore, with a lower average CBP). **B)** Considering these observations, we used the previous model (Figure 5D) as a baseline and tested what happened if we progressively reduced contralateral CBP in patients with tremor on the reference side and, if we progressively reduced ipsilateral CBP in patients without tremor on the reference side This analysis revealed that as the asymmetries increase there is a progressive increase in ipsilateral correlations and a decrease in contralateral correlations. These results are aligned with what we observed in the full dataset (Figure 5C).
